# Chronodisruption during Pregnancy Mediates the Relationship between Social Disadvantage and Reduced Brain Maturation in Neonates

**DOI:** 10.1101/2022.05.10.22274915

**Authors:** Caroline P. Hoyniak, Diana J. Whalen, Joan L. Luby, Deanna M. Barch, J. Philip Miller, Peinan Zhao, Regina L. Triplett, Yo-El Ju, Christopher D. Smyser, Barbara Warner, Cynthia E. Rogers, Erik D. Herzog, Sarah K. England

**Affiliations:** Department of Psychiatry, Washington University School of Medicine in St. Louis; Center on Biological Rhythms and Sleep, Washington University School of Medicine in St. Louis; The Program in Neuroscience, Washington University in St. Louis; Department of Psychological and Brain Sciences, Washington University in St. Louis; Department of Radiology, Washington University School of Medicine in St. Louis; Department of Biostatistics, Washington University in St. Louis; Department of Obstetrics and Gynecology, Washington University School of Medicine in St. Louis; Department of Neurology, Washington University School of Medicine in St. Louis; Department of Pediatrics, Washington University School of Medicine in St. Louis; Department of Biology, Washington University in St. Louis

**Keywords:** Adversity, Sleep, Circadian Rhythms, MRI, Infant

## Abstract

Prenatal exposure to adversity profoundly impacts offspring development. Well-documented disparities in sleep and circadian health are known to be related, and exposure to disrupted maternal sleep and circadian rhythms during pregnancy may have an effect on offspring neurodevelopment. The current study explored the association between maternal sleep and circadian rhythm disruption during pregnancy and infant brain outcomes at birth, examining sleep and circadian rhythm disruptions as a possible mediator of the effect of adversity during pregnancy on infant structural brain outcomes in 148 mother-child dyads. Maternal sleep was quantified using actigraphy data collected during each trimester of pregnancy and quantified using a measure of chronodisruption (irregularity in the sleep schedule) and a measure of chronotype (sleep timing). Adversity was quantified using a latent factor of several metrics of social disadvantage (e.g., income-to-needs ratio). Infant structural brain outcomes at birth including cortical gray matter, subcortical gray matter, and white matter volumes along with a measure of cortical folding reflecting the total surface area of the cortex. Findings indicated that chronodisruption during pregnancy was associated with smaller infant cortical gray matter, subcortical gray matter, and white matter volumes and less cortical folding at birth, with infants of mothers with later chronotypes evidencing smaller subcortical gray matter volumes. Chronodisruption during pregnancy mediated the association between maternal social disadvantage and structural brain outcomes. Findings highlight the importance of regularity and rhythmicity in sleep schedules during pregnancy and highlight the role of chronodisruption as a mechanism of the deleterious neurodevelopmental effects of prenatal adversity.

**Significance Statement:** This study examined the effect of exposure to maternal sleep and circadian rhythm disruptions during pregnancy on neonatal brain structure. Sleep and circadian disruptions were associated with global differences in neonatal brain structure. Mothers who had more irregular sleep schedules during pregnancy had infants who had smaller total cortical gray matter, subcortical gray matter, and white matter volumes and less cortical folding at birth. Irregular maternal sleep schedules during pregnancy mediated the association between adversity and structural brain outcomes, suggesting that sleep and daily rhythm disturbances may be one pathway through which adversity shapes offspring neurodevelopment. Findings imply that modifying the work schedules of pregnant women to avoid swing or night shifts might be beneficial for enhancing child neurodevelopment.

## Introduction

Early adversity has been shown to profoundly impact fetal and infant development^1^. Prenatal exposure to adversity, including poverty and exposure to stressors, trauma, abuse, and racism, is implicated in a host of negative biological, emotional, and social consequences for offspring beginning early in life and often lasting into adulthood^2,3,4,5^. A recent study demonstrated that exposure to adversity *in utero* impacts neonatal brain structure at birth^6^, with other findings showing impacts on brain structure and function in the first months of life^7,8^. Yet, despite decades of human and animal research, the pathways through which prenatal exposure to adversity shapes fetal and infant neurodevelopment remains poorly understood.

Exposure to poverty *in utero*, has been associated with enduring negative health and neurodevelopmental outcomes in offspring^7,8,9,10^. According to the fetal origins hypothesis^11,12^, factors affecting the intrauterine environment, such as poor maternal nutrition, toxin exposure, and psychosocial stress, known to be elevated in individuals exposed to poverty^13^, may have a robust effect on fetal neurodevelopment^14^. The proposed mechanisms through which intrauterine exposure to poverty affects offspring neurodevelopment include hypothalamic-pituitary-adrenal axis dysregulation and epigenetic effects^1,15^. One pathway that has received less attention is that of maternal sleep and circadian function. Sleep and circadian rhythms are intertwined but have separable mechanisms: circadian mechanisms optimally time physiological functions (including sleep, hormone secretion, and metabolism) to the 24-hour day, whereas sleep is a brain state necessary for rest and restoration that can occur at any time but is of best quality when aligned with the circadian clock^16^. There are well-documented disparities in sleep and circadian health, with individuals living in poverty more likely to have shorter sleep durations, poorer quality sleep, and to experience chronodisruption, potentially due to challenging work schedules (e.g., shift work) or increased noise and light in lower-resourced environments^17,18,19,20^. Pregnant women in poverty may be especially likely to experience sleep disturbances^21,22^, which may affect fetal neurodevelopment. Evidence across animal models and humans suggests that circadian rhythms synchronize between mother and fetus, likely through hormonal signaling via melatonin, dopamine, and corticosterone, which can cross the placenta and bind to receptors on fetal tissue (see ^23^ and ^24^ for review). Such synchronization of maternal and fetal circadian rhythms may underlie the demonstrated negative effects of irregular maternal sleep-wake cycles on fetal neurodevelopment^25^.

A growing literature suggests that, in rodents, exposure to sleep or circadian disruption in the intrauterine environment programs a range of endocrine, circadian, mood, and metabolic effects that can last into adulthood^26,27,28,29^. Sleep disruption in pregnant dams, including REM sleep deprivation and sleep restriction, is associated with brain immaturity in pups at birth, including reduced hippocampal neurogenesis^30^ and delayed maturation of neuronal groups involved in sleep and wakefulness processes^31,32^. Substantially less research has examined the consequences of prenatal maternal sleep or circadian disruptions in humans. Research suggests that human infants whose mothers had sleep disturbances during pregnancy have altered auditory event-related potentials to emotional pseudo-word stimuli at birth^33^ and show increased irritability at one month of age^34^. However, to our knowledge, no research has examined how sleep or circadian function during pregnancy affects structural neurodevelopment of offspring at birth.

In the current study, we used the longitudinal Early Life Adversity and Biological Embedding (eLABE) sample to explore the association between maternal sleep and circadian rhythm disruption during pregnancy and infant brain outcomes, both directly and as a possible mediator of the effect of exposure to adversity *in utero* on infant structural brain outcomes. To quantify sleep and circadian function during pregnancy, the current study uses two metrics of circadian function derived from actigraphy data (a measure of sleep/wake states based on wrist movements): (1) a metric of chronodisruption, which reflects irregularity in the sleep period as quantified by the standard deviation of sleep durations across the multi-week recording period, and (2) a metric of chronotype, a reflection of circadian *phase* (i.e., how early or late the daily rhythm is “set” relative to the external world) quantified by sleep period midpoint.

Previous work in this sample found that exposure to early adversity was associated with reduced infant brain volumes and cortical folding at birth^6^, including reduced white matter volume, cortical gray matter, cortical folding, and subcortical gray mater. In the current study, we focused explicitly on the tissue types and surface area metrics identified in Triplett et al. as associated with early adversity, exploring how maternal chronodisruption and chronotype during pregnancy affects these brain metrics at birth. In the current study, exposure to adversity during pregnancy was quantified by the construct of maternal social disadvantage during pregnancy, a latent factor that combined several indices of socioeconomic status and related factors, as reported by mothers during pregnancy. We test the specific hypotheses that: (1) social disadvantage during pregnancy is associated with increased chronodisruption and later chronotypes; (2) increased chronodisruption and later chronotypes is associated with smaller infant brain volumes (cortical and subcortical gray, white matter) at birth and a smaller surface area of the cortex (i.e., cortical folding); and (3) chronodisruption and chronotype mediate the association between social disadvantage during pregnancy and infant brain structure (volumes and cortical folding) at birth.

## Results

Descriptive statistics and bivariate correlations for all study variables are included in Table 1. Maternal race and social disadvantage, a latent factor combining household income-to-need ratio, neighborhood sociodemographic risk, maternal education, nutrition, and health insurance status^6^, were highly related in this sample (*M*_black_ = 1.06; *SD* = .68; *M*_white_ = -.48; *SD* = .56; *t*_140_= 14.60; *p* <. 001). As race, a socially-defined construct that is likely a proxy variable for constructs related to social disadvantage and discrimination experiences, did not offer any additional improvement to the model after the other variables were accounted for, all analyses focused on the factor of social disadvantage.

**Table 1.** Descriptive statistics and correlations: Social disadvantage, maternal chronodisruption during pregnancy, infant brain volumes at birth, and key covariates

|  | *Mean* | *Standard*<br>*Deviation* | 1 | 2 | 3 | 4 | 5 | 6 | 7 | 8 | 9 | 10 | 11 |
| --- | --- | --- | --- | --- | --- | --- | --- | --- | --- | --- | --- | --- | --- |
| 1. Social disadvantage factor score | .33 | 1.00 |  |  |  |  |  |  |  |  |  |  |  |
| 2. Chronodisruption (minutes) | 87.63 | 43.44 | .35** |  |  |  |  |  |  |  |  |  |  |
| 3. Chronotype (minutes since noon) | 900.05 | 80.51 | .54** | .26** |  |  |  |  |  |  |  |  |  |
| 4. Infant Cortical Gray Matter Volume (mm<sup>3</sup>) | 121675.80 | 15070.89 | -.36** | -.34** | -.23** |  |  |  |  |  |  |  |  |
| 5. Infant Subcortical Gray Matter Volume (mm<sup>3</sup>) | 28068.44 | 2895.03 | -.42** | -.30** | -.29** | .89** |  |  |  |  |  |  |  |
| 6. Infant White Matter Volume (mm<sup>3</sup>) | 185862.11 | 20268.36 | -.48** | -.32** | -.25** | .83** | .79** |  |  |  |  |  |  |
| 7. Cortical Folding (mm<sup>2</sup>) | 88797.95 | 11103.07 | -.44** | -.33** | -.25** | .92** | .86** | .87** |  |  |  |  |  |
| 8. Post-menstrual Age (weeks) | 41.68 | 1.33 | -.20* | -.23** | -.12 | .64** | .64** | .34** | .60** |  |  |  |  |
| 9. Infant Sex (n, % female) | 82 | 55.4 | -.07 | -.04 | -.08 | .35** | .35** | .37** | .34** | .08 |  |  |  |
| 10. Maternal Age (years) | 29.82 | 5.47 | -.52** | -.15 | -.49** | .10 | .11 | .20* | .16 | -.04 | -.12 |  |  |
| 11. Maternal Obesity (BMI) | 28.06 | 7.13 | .20* | .003 | .03 | .06 | .03 | .01 | .04 | .05 | .03 | .12 |  |
| 12. Parity (n, % first born) | 70 | 47.3 | .32** | -.001 | -.10 | -.17* | -.21* | -.23** | -.18* | -.12 | -.03 | .15 | .14 |

Note. \*\* $p < .01$ ; \* $p < .05$

### Research Question 1: Is social disadvantage associated with maternal chronodisruption and chronotype?

Linear regressions between social disadvantage and maternal chronodisruption and chronotype during pregnancy controlling for postmenstrual age at the time of the scan, infant sex, maternal age at birth, maternal obesity, and parity are presented in Table 2. Social disadvantage was associated with greater chronodisruption and later chronotype during pregnancy. Both of these relationships survived FDR correction.

**Table 2.**
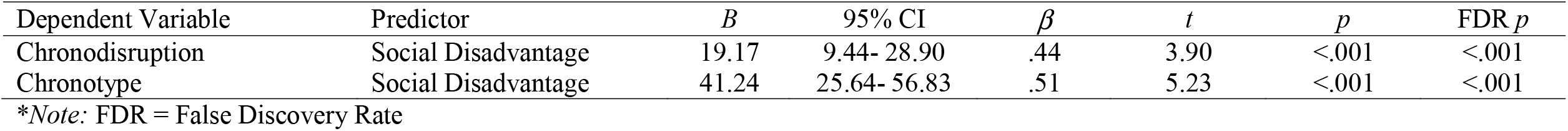
Linear regressions between social disadvantage and maternal chronodisruption controlling for postmenstrual age, infant sex, maternal age at birth, maternal obesity, and parity

| Dependent Variable | Predictor | *B* | 95% CI | $\beta$ | *t* | *p* | FDR *p* |
| --- | --- | --- | --- | --- | --- | --- | --- |
| Chronodisruption | Social Disadvantage | 19.17 | 9.44- 28.90 | .44 | 3.90 | <.001 | <.001 |
| Chronotype | Social Disadvantage | 41.24 | 25.64- 56.83 | .51 | 5.23 | <.001 | <.001 |

\*Note: FDR = False Discovery Rate

### Research Question 2: Is maternal chronodisruption or chronotype during pregnancy associated with infant brain volume at birth?

Linear regressions between maternal chronodisruption and chronotype during pregnancy and infant brain volumes at birth controlling for postmenstrual age at the time of the scan, infant sex, maternal age at birth, maternal obesity, and parity are presented in Table 3. At birth, infants of mothers who experienced more chronodisruption during pregnancy evidenced smaller total cortical gray matter, subcortical gray matter, and white matter volumes, and less cortical folding. At birth, infants of mothers who had later chronotypes during pregnancy evidenced smaller subcortical gray matter volumes. All of these relationships survived FDR correction. There were no significant associations between later sleep midpoints and infant cortical gray matter or white matter volumes, or cortical folding.

**Table 3.**
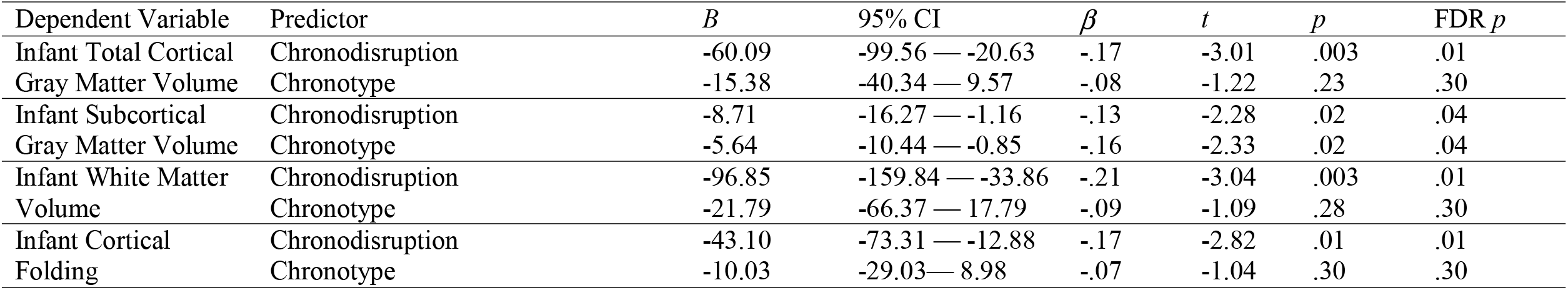
Linear regressions between maternal chronodisruption and infant brain volume at birth controlling for postmenstrual age, infant sex, maternal age at birth, maternal obesity, and parity

| Dependent Variable | Predictor | *B* | 95% CI | $\beta$ | *t* | *p* | FDR *p* |
| --- | --- | --- | --- | --- | --- | --- | --- |
| Infant Total Cortical Gray Matter Volume | Chronodisruption | -60.09 | -99.56 — -20.63 | -.17 | -3.01 | .003 | .01 |
|  | Chronotype | -15.38 | -40.34 — 9.57 | -.08 | -1.22 | .23 | .30 |
| Infant Subcortical Gray Matter Volume | Chronodisruption | -8.71 | -16.27 — -1.16 | -.13 | -2.28 | .02 | .04 |
|  | Chronotype | -5.64 | -10.44 — -0.85 | -.16 | -2.33 | .02 | .04 |
| Infant White Matter Volume | Chronodisruption | -96.85 | -159.84 — -33.86 | -.21 | -3.04 | .003 | .01 |
|  | Chronotype | -21.79 | -66.37 — 17.79 | -.09 | -1.09 | .28 | .30 |
| Infant Cortical Folding | Chronodisruption | -43.10 | -73.31 — -12.88 | -.17 | -2.82 | .01 | .01 |
|  | Chronotype | -10.03 | -29.03 — 8.98 | -.07 | -1.04 | .30 | .30 |

### Research Question 3: Does maternal chronodisruption or chronotype during pregnancy mediate associations between social disadvantage and infant brain volume at birth?

#### Chronodisruption

Four mediation models were examined to determine whether maternal social disadvantage predicted each index of infant brain volume (i.e., cortical gray matter, subcortical gray matter, white matter volumes, and cortical folding, separately) through its effect on daily deviations in sleep duration during pregnancy. Three of these four models provided evidence of significant mediation. All models controlled for postmenstrual age at the time of the scan, infant sex, maternal age at birth, maternal obesity, and parity.

Maternal social disadvantage indirectly predicted both cortical gray matter volumes and white matter through its effect on chronodisruption during pregnancy. Mothers with greater social disadvantage evidenced increased chronodisruption when compared to mothers with less social disadvantage, and infants whose mothers evidenced increased chronodisruption during pregnancy had smaller cortical gray matter (see Figure 1) and white matter volumes (see Figure 2). A bootstrap confidence interval for the indirect effects for each model based on 5,000 bootstrap samples was entirely below zero, indicating significant mediation (Figures 1 and 2). Chronodisruption accounted for 33% and 18% of the total effect of maternal social disadvantage on infant cortical gray matter volume and white matter volume, respectively.

**Figure 1.**
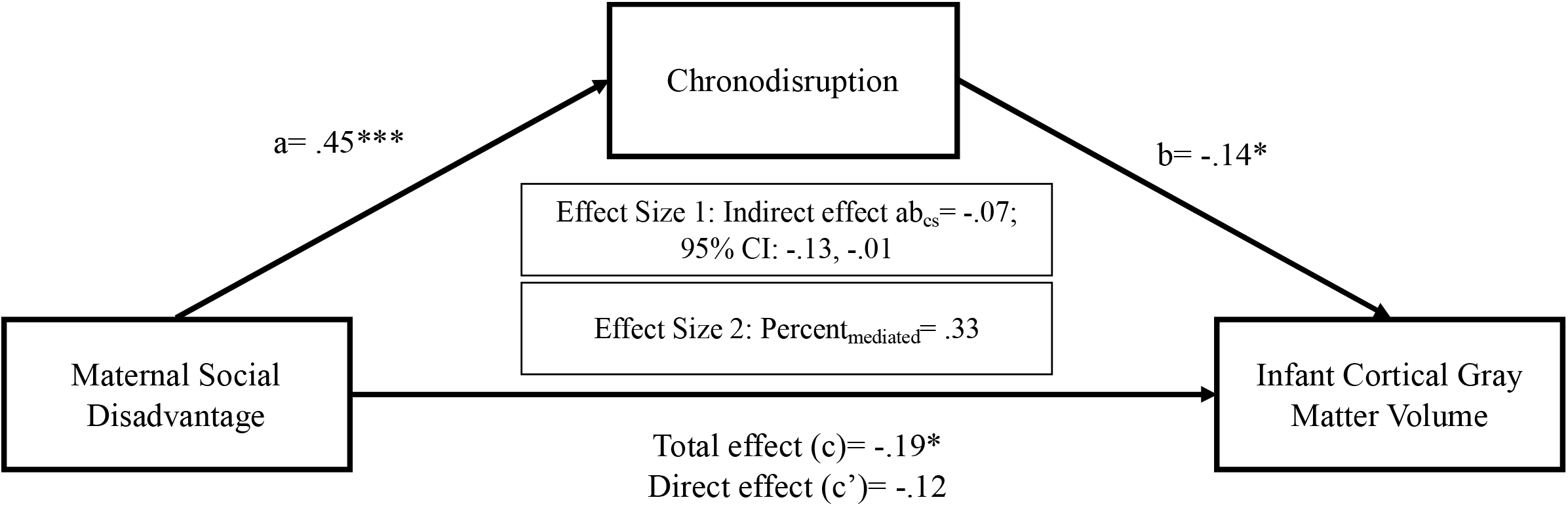
Chronodisruption mediating relationship between maternal social disadvantage and cortical gray matter volume at birth. Effect sizes of social disadvantage on chronodisruption (a) and of chronodisruption on infant cortical gray matter (b) were significant. \*\*\**p*<.001; ** *p*<.01; \**p*<.05

**Figure 2.**
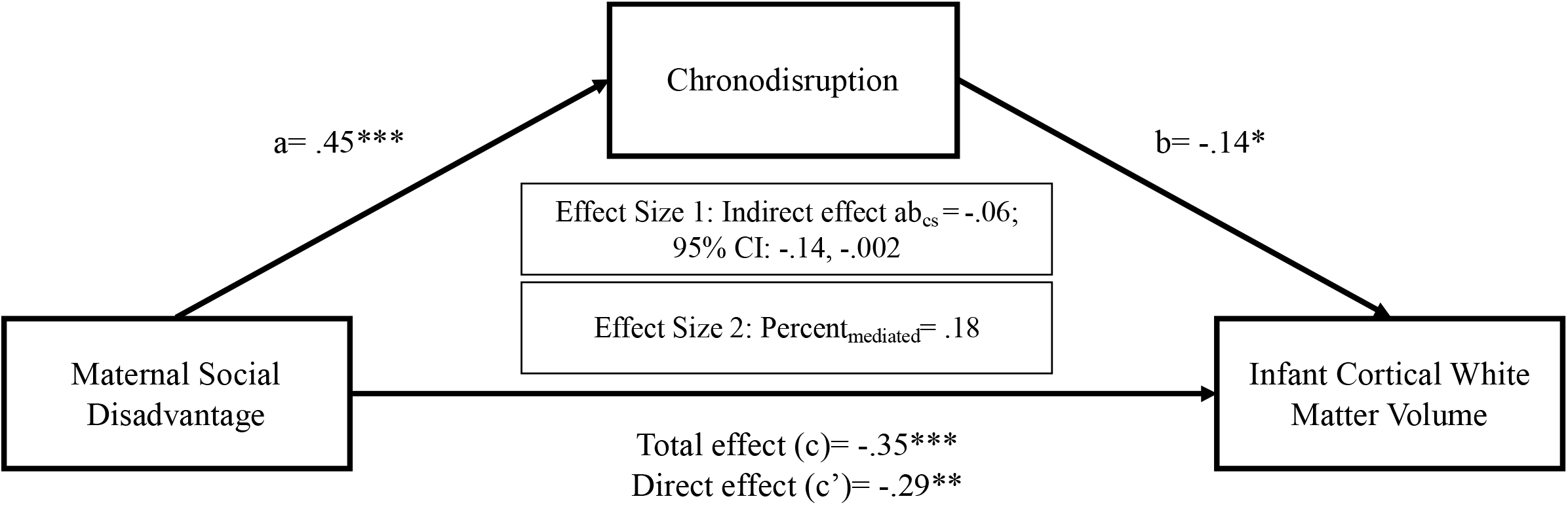
Chronodisruption mediating relationship between maternal social disadvantage and cortical white matter volume at birth. Effect sizes of social disadvantage on chronodisruption (a) and of chronodisruption on infant cortical white matter (b) were significant. \*\*\**p*<.001; ** *p*<.01; \**p*<.05

Additionally, maternal social disadvantage also indirectly predicted child cortical folding through its effect on maternal chronodisruption during pregnancy. As shown in Figure 3, mothers with greater social disadvantage evidenced increased chronodisruption during pregnancy when compared to mothers with less social disadvantage, and infants whose mothers evidenced increased chronodisruption during pregnancy had less cortical folding. The bootstrap confidence interval for the indirect effect was again entirely below zero, indicating significant mediation, with chronodisruption accounting for 18% of the total effect of maternal social disadvantage on infant cortical folding.

**Figure 3.**
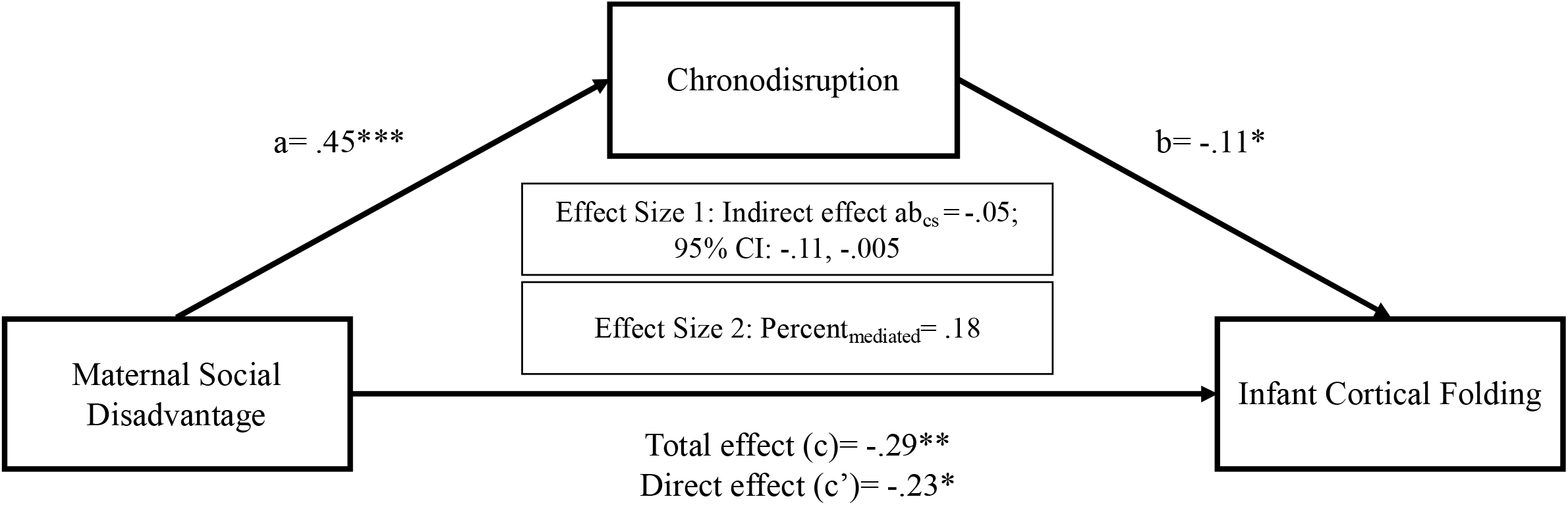
Chronodisruption mediating relationship between maternal social disadvantage and cortical folding at birth. Effect sizes of social disadvantage on chronodisruption (a) and of chronodisruption on infant cortical folding (b) were significant. \*\*\**p*<.001; ** *p*<.01; \**p*<.05

There was no evidence for significant mediation of the association between social disadvantage and infant subcortical gray matter volume by chronodisruption (see Supplemental Table S1).

#### Chronotype

Four mediation models were examined to determine whether maternal social disadvantage predicted each index of infant brain volume (i.e., cortical gray matter volumes, subcortical gray matter volumes, white matter volumes, and cortical folding) through its effect on chronotype during pregnancy. None of these models provided evidence of significant mediation (see Supplemental Table S1), suggesting chronotype does not mediate the effect of social disadvantage on infant structural brain outcomes. Of note, all models controlled for postmenstrual age at the time of the scan, infant sex, maternal age at birth, maternal obesity, and parity.

## Discussion

This study sought to test effects of maternal chronodisruption and chronotype during pregnancy on infant structural brain outcomes at birth, further exploring whether maternal daily rhythms mediate the recently described association between social disadvantage during pregnancy and structural brain alterations at birth. We found direct associations between both maternal chronodisruption and chronotype during pregnancy and infant brain structure at birth. We also found that maternal chronodisruption during pregnancy mediated the association between social disadvantage and cortical gray matter volumes, white matter volumes, and cortical folding. These findings suggest that maternal chronodisruption during pregnancy is a pathway through which social disadvantage leads to disrupted infant neural development.

Our results add to an emerging literature on the association between adversity/disadvantage and sleep and chronodisruption during pregnancy. We found that higher levels of social disadvantage are associated with greater chronodisruption and a later chronotype during pregnancy. This extends prior research suggesting an association between lower socioeconomic status and poorer perceived sleep quality in pregnant women^22^, and between poverty and heightened insomnia symptoms^21^. Increased exposure to stressors, pre-sleep worries, and environmental conditions that are non-conducive to sleep (e.g., environmental noise, light pollution, parent-child bedsharing) may underlie these associations.

We also found that greater irregularity in maternal sleep-wake patterns during pregnancy was associated with smaller infant total cortical gray matter, subcortical gray matter, and white matter volumes and less cortical folding. Further, at birth, infants of mothers with later chronotypes evidenced smaller subcortical gray matter volumes. These findings held even after accounting for postmenstrual age, infant sex, maternal age at birth, maternal obesity, and parity. An emerging literature shows that sleep/wake disruptions during pregnancy affect birth outcomes^35,36,37^, however this is the first study that we are aware of that has found relations between maternal daily rest-activity rhythms during pregnancy and infant neural outcomes. These findings have direct implications for pregnant women employed in professions that disrupt circadian function and sleep timing through requiring irregular or nighttime working hours.

There are a number of plausible mechanisms through which sleep disturbances and chronodisruption may provide an adverse intrauterine environment and affect neurodevelopmental outcomes in offspring. For example, a growing literature exploring these mechanisms highlights key roles for melatonin and cortisol, circadian-controlled hormones, in fetal neurodevelopment^23,38^. Melatonin and cortisol levels increase during pregnancy, can cross the placenta and bind to their cognate receptors in fetal tissue^24,39^. In rodents, melatonin or glucocorticoid administration can shift fetal daily rhythms and influence subsequent development of organ systems and behaviors such as sleep-wake rhythms^40,41^. Disruptions in either the daily patterning or peak levels of these hormones during pregnancy can have long term consequences on offspring development^38^. Additionally, increased chronodisruption and later chronotypes are associated with sleep disturbances, including disruptions in sleep quality and quantity^42,43^. The disruption of sleep and its restorative mechanisms that results from maternal chronodisruption/late chronotypes may also have downstream effects on offspring neurodevelopment.

We also found that chronodisruption during pregnancy, in part, explained why some infants with greater maternal social disadvantage evidence smaller cortical gray matter volume, smaller white matter volume, and less cortical folding. These findings highlight the importance of maternal sleep and daily rhythms during pregnancy for child neurodevelopmental outcomes. However, it is not entirely clear why chronodisruption mediated the relationship of social disadvantage to the cortical gray matter, cortical folding and total white matter (which is dominated by cortical white matter), but not subcortical gray matter. One possibility is that the relationship of social disadvantage to subcortical gray matter volume is more strongly influenced by other factors associated with social disadvantage, such as maternal cortisol secretion or exposure to environmental toxins. Regardless, the role of maternal daily rhythms during pregnancy on child outcomes is highly understudied, and ripe with potential for relatively low cost and brief behavioral interventions. Future studies also should examine whether the significant mediations between maternal sleep and infant brain volumes are associated with neural and behavioral outcomes later in childhood.

This study had a number of strengths and adds meaningfully to a sparse literature on the effects of prenatal chronodisruption in humans. Our mediation models had temporal precedence: prenatal social disadvantage was measured prior to and at the same time as maternal daily rhythms and both were measured prior to infant brain volumes. Our sample also included in-depth, objective sleep data collection each trimester of pregnancy, which can avoid subjective errors associated with self-report sleep questionnaires. This study also has several limitations worthy of consideration. The sample size (*n* = 148), while large for infant neuroimaging, limits our power to detect small effects. Additionally, we were not able to examine race in this analysis, give high levels of collinearity between race and social disadvantage in our sample. Although collinearity of race and social disadvantage is often characteristic of US samples, the consideration of additional constructs such as social exclusion and discrimination and how these constructs intersect with both race and social disadvantage will be crucial to future research in this sample and in this research area.

Study findings have direct implications for modifying work schedules of pregnant women whose work/routines require chronodisruption. From a public health perspective, minimizing the need for pregnant individuals to work swing or night shifts might be beneficial for both improving pregnancy outcomes and enhancing child neurodevelopment. Clinically, results from this study indicate the importance of interventions during pregnancy to improve the sleep and circadian health of mothers. Such interventions may not only enhance infant brain development and related cognitive/behavioral outcomes, they may also reduce social disadvantage-related health disparities that perpetuate across generations.

Low-cost, brief behavioral interventions provided to individuals experiencing chronodisruption during pregnancy may significantly improve infant health and wellbeing. Additional critical questions remain about the continued influence of prenatal social disadvantage and sleep/circadian disruptions on child neurodevelopment throughout infancy and early development. Many of these prenatal effects may be exacerbated, or possibly ameliorated, by aspects of the parent-child relationship, child and parent social support, exposure to ongoing adversity, etc. Future research will be critical for understanding how maternal sleep and circadian disruptions during pregnancy continue to influence child development across the first years of life and beyond.

## Materials and Methods

### Participants

Participants in the current study included 142 mothers-infant dyads enrolled in two large overlapping studies: the 1000 Women Cohort (1KWC) and the eLABE study. In the 1KWC, 1220 women with a live birth (including 679 Black and 542 low-socioeconomic status women) were assessed throughout their pregnancy, providing blood samples, detailed survey data, circadian data (actigraphy and daily hormonal profiles), and birth outcome data. Mothers were recruited during the first trimester and followed throughout pregnancy and at delivery. In the 1KWC, low-socioeconomic status was defined as having an annual salary of <$24,999 and/or being the recipient of government assistance. Exclusion criteria for the 1KWC included: multiple gestations, diagnosed infections known to cause congenital disease (e.g., syphilis), and maternal alcohol or drug use during pregnancy (excluding tobacco and marijuana). In their third trimester, a subsample (*n*= 399) of the participants in the 1KWC were recruited to participate in eLABE, an ongoing, longitudinal study of child development from birth to age 3.

Of the 399 1KWC mother-infants dyads who participated in the eLABE study, 201 provided usable actigraphy data during trimesters 1 and 2 of pregnancy. Of these, 148 full-term infants (born > 37 weeks gestational age) had high-quality magnetic resonance imaging (MRI) data and met our previously defined inclusion criteria^6^: birth weight > 2000 g, no Neonatal Intensive Care Unit admission >7 days, and no evidence of brain injury on MRI (final sample of 148 mother-infant dyads). The racial breakdown of the children in the final sample was 51% White, 47% Black or African American, 2% Other, and families had an average Income-to-Needs ratio of 3.84 (*SD*= 3.09, range= 0.44 – 12.04). Mothers ranged in age from 18.77 - 41.80 years (*M*= 29.83 years; *SD* = 5.47 years) and at the time of MRI scan, infants were an average postmenstrual age of 41.68 weeks (range 38–45 weeks). The Washington University Human Research Protection Office gave ethical approval for this work. Informed consent was obtained for each participant and subsequent parental informed consent was obtained for each infant prior to participation. Data were collected with consent forms that allow data sharing. De-identified data is being deposited in the NIMH Data Archive and will be available after the conclusion of the eLABE 3-year wave.

### Measures

#### Social Disadvantage

Adversity-related information, including health insurance status, income, maternal education, address, household composition, and maternal nutrition was obtained from participants either during pregnancy or at delivery. Confirmatory factor analysis (CFA) was used to derive a latent factor reflecting maternal social disadvantage during pregnancy (CFA results detailed in ^44^). The following indexes were used to estimate the Social Disadvantage latent factor: (1) household Income to Needs Ratio^45^ in each trimester of pregnancy, (2) national Area Deprivation Index percentiles^46^, a measure of sociodemographic disadvantage at the census block level, were calculated based on family addresses at birth, (3) maternal nutrition as measured by the Healthy Eating Index^47^ completed by mothers during the third trimester or at delivery, (4) highest level of maternal educational attainment at the time of study enrollment, and (5) health insurance status at the time of study enrollment (verified by mothers in the third trimester or at delivery). This latent factor was used in subsequent analysis. As race was highly correlated with the social disadvantage factor and offered no additional improvement to the model after other variables were accounted for, race was not included in the latent social disadvantage factor^44^.

#### Maternal Chronodisruption and Chronotype during Pregnancy

A minimum of two weeks of wrist actigraphy data were collected from mothers during their first, second, and third trimesters of pregnancy. Actigraphs contain an accelerometer that measures minute-by-minute motor activity, which allows for the estimation of sleep and wake patterns. Actigraphy watches, the MotionWatch 8 (CamNtech Ltd., Cambridge, U.K.), were worn by mothers on their non-dominant wrist. The MotionWatch 8 contains a piezoelectric accelerometer with a sensitivity greater than 0.05 g, a sampling frequency of 32 Hz, and bandpass filter of 3-11 Hz. The actigraphy data was collected in 1-minute epochs. The actigraphy data were processed using MotionWare software 2.5 (CamNtech Ltd) and analyzed using a high-throughout, automated method to derive the sleeping time indexes, such as daily sleep onset and offset times, duration and mid-time, sleeping time inter-daily variability, etc.^48^ We use two metrics of daily rhythms derived from actigraphy data: (1) a metric of chronodisruption, which reflects irregularity in the sleep period as quantified by the standard deviations of sleep durations (defined as the length of time between sleep period onset and offset times) across the recording period, and (2) a metric of chronotype, a reflection of circadian *phase* (i.e., how early or late the circadian rhythm is relative to the external world) quantified by sleep period midpoint. The sleep period midpoint was defined as the time equidistant between sleep onset time and offset times. The median values of each of these sleep indexes across the trimesters of pregnancy was calculated and used in analyses.

#### Infant Brain Volumes at Birth: MRI Acquisition, Processing, and Measures

All infants underwent non-sedated MRI scans within the first weeks of life using a Siemans Prisma 3T scanner and a 64-channel head coil. MRI sequence parameters and our standardized preprocessing pipeline are further detailed in ^6^. Briefly, in this analysis, brain volumetric measures of interest included the total white matter, cortical and subcortical gray matter, and cortical surface area. To generate these measures, the age-specific, automated, Melbourne Children’s Regional Infant Brain atlas Surface (M-CRIB-S) segmentation and surface extraction toolkit was applied to high-quality (low-motion), preprocessed T2-weighted images^49,50^. The M-CRIB-S toolkit output included spatially normalized (within group and to the M-CRIB atlas) segmentations and surface-based cortical parcellations of the white and gray matter, cerebellum, brainstem, and subcortical gray matter subdivisions, with FreeSurfer-like labeling. All segmentations and surfaces were qualitatively inspected for accuracy, manually edited as necessary, and designated as complete by a highly-experienced team of two imaging scientists and a pediatric neurologist (C.D.S.).

#### Covariates

The following covariates were included in analyses: PMA, infant sex, maternal age at birth, pre-pregnancy maternal obesity (calculated using standard procedures, Body Mass Index >=30), and parity.

### Data-Analytic Plan

Linear regressions were conducted in order to determine whether social disadvantage and/or infant brain volume at birth were associated with maternal chronodisruption during pregnancy (research questions 1 and 2). Covariates included PMA, infant sex, maternal age at birth, maternal obesity, and parity. Models were conducted separately for daily deviation in sleep duration and daily sleep midpoint, two indices of maternal chronodisruption. In line with our past work^6^, the following infant brain volume regions were examined: total cortical gray matter, subcortical gray matter, and total white matter. Cortical folding was measured using the total surface area of the cortex for both hemispheres. False discovery rate (FDR) correction was used to account for multiple comparisons in the regression analyses.

Simple mediation analyses using ordinary least squares path analyses^51^ were conducted to determine whether maternal chronodisruption during pregnancy mediated the association between social disadvantage and infant brain volumes and cortical surface area. In all models, social disadvantage across pregnancy was the predictor. Two maternal sleep indexes (daily deviation in sleep duration and daily sleep midpoint) were independently tested as mediators of the infant brain volumes found to be significant in regression analyses using the SPSS PROCESS macro^52^. Covariates (PMA, infant sex, maternal age at birth, maternal obesity, and parity) were applied to both mediators and outcome variables.

## Data Availability

For most of the data included as a part of the current study, data were collected with consent forms that allow data sharing. De-identified data is being deposited in the NIMH Data Archive and will be available after the conclusion of the eLABE 3-year wave. For the maternal sleep variables specifically: The data are not publicly available as the minimal data set for this study on pregnant participants contains identifying patient-level data which cannot be suitably de-identified or aggregated. Additionally, a subset of participants in the 1KWC did not consent for future research in the patient consent form approved by the Institutional Review Board at Washington University in St. Louis. Proposals for access to this data should be directed to, Senior Clinical Research Coordinator in the Division of Clinical Research in the Department of Obstetrics and Gynecology. To gain access, data requestors will need to sign a data access agreement.

## Acknowledgements

The authors wish to thank the mothers and infants participating in the 1KWC and eLABE studies for their time and dedication to these ongoing projects.

## References

1. Barrero-Castillero, A., Morton, S. U., Nelson, C. A. & Smith, V. C. Psychosocial Stress and Adversity: Effects from the Perinatal Period to Adulthood. Neoreviews 20, e686–e696 (2019).

2. Ahmad, S. I. et al. Maternal childhood trauma and prenatal stressors are associated with child behavioral health. J Dev Orig Hlth Dis 1–11 (2021) doi:10.1017/s2040174421000581.

3. Conradt, E., Carter, S. E. & Crowell, S. E. Biological Embedding of Chronic Stress Across Two Generations Within Marginalized Communities. Child Dev Perspect 14, 208–214 (2020).

4. Monk, C., Spicer, J. & Champagne, F. A. Linking prenatal maternal adversity to developmental outcomes in infants: The role of epigenetic pathways. Dev Psychopathol 24, 1361–1376 (2012).

5. Rakers, F. et al. Transfer of maternal psychosocial stress to the fetus. Neurosci Biobehav Rev 117, 185–197 (2017).

6. Triplett, R. L. et al. Prenatal Exposure to Early Life Adversity and Neonatal Brain Volumes at Birth. Medrxiv 2021.12.20.21268125 (2021) doi:10.1101/2021.12.20.21268125.

7. Betancourt, L. M. et al. Effect of socioeconomic status (SES) disparity on neural development in female African-American infants at age 1 month. Developmental Sci 19, 947–956 (2016).

8. Knickmeyer, R. C. et al. Impact of Demographic and Obstetric Factors on Infant Brain Volumes: A Population Neuroscience Study. Cereb Cortex 27, 5616–5625 (2016).

9. Gilman, S. E. et al. Socioeconomic disadvantage, gestational immune activity, and neurodevelopment in early childhood. Proc National Acad Sci 114, 6728–6733 (2017).

10. Johnson, S. B., Riis, J. L. & Noble, K. G. State of the Art Review: Poverty and the Developing Brain. Pediatrics 137, e20153075 (2016).

11. Almond, D. & Currie, J. Killing Me Softly: The Fetal Origins Hypothesis. J Econ Perspect 25, 153–172 (2011).

12. Barker, D. J. P. Fetal origins of coronary heart disease. Bmj 311, 171 (1995).

13. Evans, G. W. & Kantrowitz, E. SOCIOECONOMIC STATUS AND HEALTH: The Potential Role of Environmental Risk Exposure. Public Health 23, 303–331 (2002).

14. Antonelli, M. C., Pallarés, M. E., Ceccatelli, S. & Spulber, S. Long-term consequences of prenatal stress and neurotoxicants exposure on neurodevelopment. Prog Neurobiol 155, 21–35 (2017).

15. Scorza, P. et al. Research Review: Intergenerational transmission of disadvantage: epigenetics and parents’ childhoods as the first exposure. J Child Psychol Psyc 60, 119–132 (2019).

16. Chronobiology of Sleep. in Encyclopedia of Sleep vol. 4 407–445 (Elsevier, Inc., 2013).

17. Billings, M. E. et al. Disparities in Sleep Health and Potential Intervention Models. Chest 159, 1232–1240 (2021).

18. Mezick, E. J. et al. Influence of Race and Socioeconomic Status on Sleep: Pittsburgh SleepSCORE Project. Psychosom Med 70, 410–416 (2008).

19. Patel, N. P., Grandner, M. A., Xie, D., Branas, C. C. & Gooneratne, N. “Sleep disparity” in the population: poor sleep quality is strongly associated with poverty and ethnicity. Bmc Public Health 10, 475 (2010).

20. Johnson, D. A., Billings, M. E. & Hale, L. Environmental Determinants of Insufficient Sleep and Sleep Disorders: Implications for Population Health. Curr Epidemiology Reports 5, 61–69 (2018).

21. Kalmbach, D. A. et al. Insomnia, Short Sleep, And Snoring In Mid-To-Late Pregnancy: Disparities Related To Poverty, Race, And Obesity. Nat Sci Sleep 11, 301–315 (2019).

22. Okun, M. L., Tolge, M. & Hall, M. Low Socioeconomic Status Negatively Affects Sleep in Pregnant Women. J Obstetric Gynecol Neonatal Nurs 43, 160–167 (2014).

23. Bates, K. & Herzog, E. D. Maternal-Fetal Circadian Communication During Pregnancy. Front Endocrinol 11, 198 (2020).

24. Mark, P. J., Crew, R. C., Wharfe, M. D. & Waddell, B. J. Rhythmic Three-Part Harmony: The Complex Interaction of Maternal, Placental and Fetal Circadian Systems. J Biol Rhythm 32, 534–549 (2017).

25. Pires, G. N. et al. Effects of sleep modulation during pregnancy in the mother and offspring: Evidences from preclinical research. J Sleep Res 30, e13135 (2021).

26. Varcoe, T. J., Gatford, K. L. & Kennaway, D. J. Maternal circadian rhythms and the programming of adult health and disease. Am J Physiology-regulatory Integr Comp Physiology 314, R231–R241 (2018).

27. Varcoe, T. J., Wight, N., Voultsios, A., Salkeld, M. D. & Kennaway, D. J. Chronic Phase Shifts of the Photoperiod throughout Pregnancy Programs Glucose Intolerance and Insulin Resistance in the Rat. Plos One 6, e18504 (2011).

28. Mendez, N. et al. Gestational Chronodisruption Impairs Circadian Physiology in Rat Male Offspring, Increasing the Risk of Chronic Disease. Endocrinology 157, 4654–4668 (2016).

29. Smarr, B. L., Grant, A. D., Perez, L., Zucker, I. & Kriegsfeld, L. J. Maternal and Early-Life Circadian Disruption Have Long-Lasting Negative Consequences on Offspring Development and Adult Behavior in Mice. Sci Rep-uk 7, 3326 (2017).

30. Zhao, Q. et al. Maternal sleep deprivation inhibits hippocampal neurogenesis associated with inflammatory response in young offspring rats. Neurobiol Dis 68, 57–65 (2014).

31. Aswathy, B. S., Kumar, V. M. & Gulia, K. K. Immature sleep pattern in newborn rats when dams encountered sleep restriction during pregnancy. Int J Dev Neurosci 69, 60–67 (2018).

32. Aswathy, B. S., Kumar, V. M. & Gulia, K. K. The effects of rapid eye movement sleep deprivation during late pregnancy on newborns’ sleep. J Sleep Res 27, 197–205 (2018).

33. Lavonius, M. et al. Maternal sleep quality during pregnancy is associated with neonatal auditory ERPs. Sci Rep-uk 10, 7228 (2020).

34. Nakahara, K. et al. Association of maternal sleep before and during pregnancy with preterm birth and early infant sleep and temperament. Sci Rep-uk 10, 11084 (2020).

35. Facco, F. L. et al. Later sleep timing is associated with an increased risk of preterm birth in nulliparous women. Am J Obstetrics Gynecol Mfm 1, 100040 (2019).

36. Warland, J., Dorrian, J., Morrison, J. L. & O’Brien, L. M. Maternal sleep during pregnancy and poor fetal outcomes: A scoping review of the literature with meta-analysis. Sleep Med Rev 41, 197–219 (2018).

37. Reschke, L. et al. Chronodisruption: An Untimely Cause of Preterm Birth? Best Pract Res Cl Ob 52, 60–67 (2018).

38. Sagrillo-Fagundes, L., Salustiano, E. M. A., Yen, P. W., Soliman, A. & Vaillancourt, C. Melatonin in Pregnancy: Effects on Brain Development and CNS Programming Disorders. Curr Pharm Design 22, 978–986 (2016).

39. Reppert, S. M., Weaver, D. R., Rivkees, S. A. & Stopa, E. G. Putative Melatonin Receptors in a Human Biological Clock. Science 242, 78–81 (1988).

40. Davis, F. C. & Mannion, J. Entrainment of hamster pup circadian rhythms by prenatal melatonin injections to the mother. Am J Physiology-regulatory Integr Comp Physiology 255, R439–R448 (1988).

41. Cecmanová, V., Houdek, P., Šuchmanová, K., Sládek, M. & Sumová, A. Development and Entrainment of the Fetal Clock in the Suprachiasmatic Nuclei: The Role of Glucocorticoids. J Biol Rhythm 34, 307–322 (2019).

42. Vetter, C. Circadian disruption: What do we actually mean? Eur J Neurosci 51, 531–550 (2020).

43. Selvi, Y. et al. Comparison of dream anxiety and subjective sleep quality between chronotypes. Sleep Biol Rhythms 10, 14–22 (2012).

44. Luby, J. L. et al. Modeling Prenatal Adversity/Advantage: Effects on Birth Weight. Medrxiv 2021.12.16.21267938 (2021) doi:10.1101/2021.12.16.21267938.

45. Brooks-Gunn, J. & Duncan, G. J. The Effects of Poverty on Children. Futur Child 7, 55 (1997).

46. Kind, A. J. H. & Buckingham, W. R. Making Neighborhood-Disadvantage Metrics Accessible — The Neighborhood Atlas. New Engl J Med 378, 2456–2458 (2018).

47. Krebs-Smith, S. M. et al. Update of the Healthy Eating Index: HEI-2015. J Acad Nutr Diet 118, 1591–1602 (2018).

48. Zhao, P. Sleep behavior and chronotype before and throughout pregnancy. Sleep Medicine.

49. Adamson, C. L. et al. Parcellation of the neonatal cortex using Surface-based Melbourne Children’s Regional Infant Brain atlases (M-CRIB-S). Sci Rep-uk 10, 4359 (2020).

50. Alexander, B. et al. A new neonatal cortical and subcortical brain atlas: the Melbourne Children’s Regional Infant Brain (M-CRIB) atlas. Neuroimage 147, 841–851 (2017).

51. Hayes, A. F. Introduction to mediation, moderation, and conditional process analysis: A regression-based approach. (Guilford Press, 2013).

52. Hayes, A. F. PROCESS: A Versatile Computational Tool for Observed Variable Mediation, Moderation, and Conditional Process Modeling. (2012).

